# Trajectory Modeling and Response Prediction in Transcranial Magnetic Stimulation for Depression

**DOI:** 10.1101/2024.05.30.24308258

**Authors:** Aaron N. McInnes, Sarah T. Olsen, Christi R.P. Sullivan, Dawson C. Cooper, Saydra Wilson, Ayse Irem Sonmez, Sophia C. Albott, Stephen C. Olson, Carol B. Peterson, Barry R. Rittberg, Alexander Herman, Matej Bajzer, Ziad Nahas, Alik S. Widge

## Abstract

Repetitive transcranial magnetic stimulation (rTMS) therapy could be improved by better and earlier prediction of response. Latent class mixture (LCMM) and non-linear mixed effects (NLME) modelling have been applied to model the trajectories of antidepressant response (or non-response) to TMS, but it is not known whether such models can predict clinical outcomes. We compared LCMM and NLME approaches to model the antidepressant response to TMS in a naturalistic sample of 238 patients receiving rTMS for treatment resistant depression (TRD), across multiple coils and protocols. We then compared the predictive power of those models. LCMM trajectories were influenced largely by baseline symptom severity, but baseline symptoms provided little predictive power for later antidepressant response. Rather, the optimal LCMM model was a nonlinear two-class model that accounted for baseline symptoms. This model accurately predicted patient response at 4 weeks of treatment (AUC = 0.70, 95% CI = [0.52-0.87]), but not before. NLME offered slightly improved predictive performance at 4 weeks of treatment (AUC = 0.76, 95% CI = [0.58 – 0.94], but likewise, not before. In showing the predictive validity of these approaches to model response trajectories to rTMS, we provided preliminary evidence that trajectory modeling could be used to guide future treatment decisions.

## 1. Introduction

Repetitive transcranial magnetic stimulation (rTMS) of the prefrontal cortex (PFC) is effective in treatment resistant depression (TRD) (McClintock et al., 2018; O’Reardon et al., 2007). However, response to rTMS varies greatly (Kaster et al., 2020, 2023; Schilberg et al., 2017), with a 40-50% non-response rate in clinical practice (Sackeim et al., 2020; Taylor et al., 2017). Furthermore, a typical course of rTMS is 5 days a week for 4-6 weeks (Sackeim et al., 2020), a significant time burden for patients. Novel protocols offer the possibility of more rapid response (Cole et al., 2020), but require even greater time investment and clinical staffing. At the same time, for many centers, demand outstrips the available treatment slots, leading to waiting lists. Those lists might be shortened if treatment could be stopped (or altered) earlier for non-responders. Therefore, there is a need to understand who will respond to rTMS and when (Baeken et al., 2019). Modeling inter-individual variability may allow for earlier detection of treatment (non)response, and eventually, more personalized treatment (Baeken et al., 2019).

A critical first step is moving beyond response at a single post-treatment timepoint, and recognizing that clinical response evolves over several weeks in stereotypical patterns. Latent class mixture modeling (LCMM), or growth mixture modeling (GMM) (Muthén & Asparouhov, 2008; Muthén & Shedden, 1999) has been used to model the trajectories of depression response to treatments: pharmacological (Smagula et al., 2015; Uher et al., 2010), pharmacological plus psychotherapy (Stulz et al., 2010), and rTMS (Kaster et al., 2019, 2020, 2023). LCMM and GMM identify “latent classes”: groups of patients whose response trajectories are similar and can be captured by a simple function. Trajectory modeling approaches could allow early discrimination of individuals who are on paths leading to (non)response (Uher et al., 2010).

Two studies from the same group have identified multiple trajectories of response to rTMS for depression (Kaster et al., 2020, 2019). Non-response trajectories were identified in both studies, and non-responders had higher baseline symptoms than patients on response trajectories. Both studies identified various linear response trajectories, differing in baseline symptoms (intercept) or level of improvement (slope), as well as a rapid response trajectory in one model (Kaster et al., 2019). Response rates differed among the trajectories at week 3 in the absence of a rapid response group (Kaster et al., 2020), and as early as week 1 with rapid responders (Kaster et al., 2019).

However, neither of these studies analyzed whether the trajectories could predict future treatment response during a TMS treatment course. Further, modelling was run with clinical trial data, which had much stricter exclusion criteria and control of treatment protocols than in typical clinical practice. It is unknown whether these trajectories generalize to a more heterogeneous, naturalistic sample. Finally, these studies did not cross-validate their models’ predictive performance. Cross-validation is a standard method for assessing how well a model might generalize to new patients (Cawley & Talbot, 2010) and has been repeatedly identified as necessary for biomarker studies (Grzenda et al., 2021; Poldrack et al., 2020; Widge et al., 2019).

As an alternative approach, Berlow et al. (2023) modelled response trajectories using nonlinear mixed effects (NLME) models which treat symptom response to TMS as an exponential decay of symptom severity over the course of treatment. That study, which did cross-validate, reported high sensitivity/specificity for response prediction as early as the first week of treatment. If replicated, that capacity for early prediction could substantially alter TMS treatment practices.

Here, we perform an independent predictive validation of trajectory modeling in TMS, comparing both approaches above. We used LCMM/NLME to model response trajectories in a naturalistic sample containing multiple coils and treatment protocols. For each approach, we identified the optimal level of model complexity, which in part replicate and in part diverge from prior studies. Given the optimal models, we tested and compared the clinical predictive power of each method, by predicting response of a held-out sample using the first few weeks of clinical data.

## 2. Methods

### 2.1. Sample

Data were derived from a registry of 238 patients receiving rTMS for TRD at a clinic in Minnesota. If a patient completed more than one series of rTMS, only the data from their first treatment series was used. Retrospective review of patient data related to their TMS treatment was approved by the University of Minnesota Institutional Review Board. See Table 1 for demographic information.

**Table 1.** Patient characteristics for the whole sample, and for each of the identified trajectories. *Race excludes 15 patients for whom there was no race recorded. **Responder is defined as >50% reduction in PHQ-9 after 6 weeks of treatment. ***ICD Diagnosis excludes 24 patients for whom there were no diagnoses recorded. ****Treatment year is based on the year the clinic began treating patients.

|  |  | All Patients | Assigned Improvement Trajectory | Assigned Non-Response Trajectory |
| --- | --- | --- | --- | --- |
| Trajectory Modeling Data | n | 238 | 97 | 141 |
|  | % of total sample | 100 | 40.76 | 59.24 |
|  | Posterior Probability | NA | 0.89 | 0.84 |
| Demographics | Age- M (SD) | 44.90 (15.26) | 45.45 (14.32) | 44.52 (15.91) |
|  | Sex- n males (%) | 87 (36.55) | 34 (35.05) | 53 (37.59) |
|  | *Race- n minority (%) | 18 (8.07) | 8 (8.25) | 10 (7.09) |
|  | Antianxiety use - n yes (%) | 144 (60.5) | 56 (57.73) | 88 (62.41) |
|  | Antidepressant use – n yes (%) | 225 (94.54) | 94 (96.91) | 131 (92.91) |
|  | Antipsychotic use – n yes (%) | 115 (48.32) | 47 48.45) | 68 (48.23) |
|  | Stimulant use - n yes (%) | 119 (50) | 46 (47.42) | 73 (51.77) |
|  | Other psychotherapeutic use – n yes (%) | 19 (7.98) | 6 (6.19) | 13 (9.22) |
|  | Number of treatments- M (SD) | 57.35 (19.71) | 56.74 (12.54) | 57.77 (23.44) |
|  | Baseline PHQ9- M (SD) | 17.86 (4.56) | 18.01 (4.47) | 17.76 (4.63) |
|  | Final PHQ9- M (SD) | 12.76 (6.58) | 11.14 (5.75) | 15.71 (5.37) |
|  | **Responder- n (%) | 44 (18.49) | 40 (41.24) | 4 (2.84) |
| ICD-10 Diagnosis*** | Disorders due to known physiological conditions (F01-F09) - n (%) | 3 (1.4) | 0 (0) | 3 (2.13) |
|  | Substance use disorders (F10-F19) - n (%) | 36 (16.82) | 14 (14.43) | 22 (15.6) |
|  | Primary psychoses (F20-F29) - n (%) | 2 (0.93) | 0 (0) | 2 (1.42) |
|  | Mood disorders (F30-F39) – n (%) | 214 (100) | 88 (100) | 126 (100) |
|  | Anxiety, stress, and somatoform disorders (F40-F48) – n (%) | 128 (59.81) | 54 (55.67) | 74 (52.48) |
|  | Behavioral disorders with physical factors (F50-F59) – n (%) | 41 (19.16) | 16 (16.49) | 25 (17.73) |
|  | Personality and behavioral disorders (F60-F69) – n (%) | 24 (11.21) | 14 (14.43) | 10 (7.09) |
|  | Intellectual disabilities (F70-F79) – n (%) | 0 (0) | 0 (0) | 0 (0) |
|  | Developmental disorders (F80-F89) – n (%) | 7 (3.27) | 5 (5.15) | 2 (1.42) |
|  | Behavioral and emotional disorders with onset in childhood (F90-F98) – n (%) | 41 (19.16) | 14 (14.43) | 27 (19.15) |
|  | Total comorbid with mood disorders (F30-F39) - n (%) | 153 (71.5) | 64 (65.98) | 89 (63.12) |
| Treatment Year**** | Year 1- n (%) | 57 (23.95) | 27 (27.84) | 30 (21.28) |
|  | Year 2- n (%) | 72 (30.25) | 30 (30.93) | 42 (29.79) |
|  | Year 3- n (%) | 60 (25.21) | 21 (21.65) | 39 (27.66) |
|  | Year 4- n (%) | 40 (16.81) | 14 (14.43) | 26 (18.44) |
|  | Year 5- n (%) | 9 (3.78) | 5 (5.15) | 4 (2.84) |
| Treatment coil | F8- n (%) | 144 (60.50) | 53 (54.64) | 91 (64.54) |
|  | H1- n (%) | 71 (29.83) | 36 (37.11) | 35 (24.82) |
|  | Switch- n (%) | 23 (9.66) | 8 (8.25) | 15 (10.64) |
| Treatment protocol | High frequency- n (%) | 83 (34.87) | 41 (42.27) | 42 (29.79) |
|  | Low frequency- n (%) | 12 (5.04) | 6 (6.19) | 6 (4.26) |
|  | iTBS- n (%) | 104 (43.70) | 40 (41.24) | 64 (45.39) |
|  | Switch- n (%) | 39 (16.39) | 10 (10.31) | 29 (20.57) |

Patients received TMS using either a Brainsway H1 coil (H1; n = 71) or Magstim air-cooled figure of 8 coil (Horizon® Performance and Horizon® Light; F8; n = 144) targeted by the Beam-F3 method (Beam et al., 2009). Some patients also switched coils during their treatment (n = 23). Among those who did not switch coils, we assessed for differences in demographic characteristics between those receiving H1 or F8 treatment (using linear regression for numeric variables, and logistic regression for binary variables).

F8 treatment included 10 Hz (n = 10; high frequency), 1 Hz (n = 12; low frequency), and intermittent theta burst stimulation (iTBS, n = 104) protocols. H1 treatment used an 18 Hz protocol (n = 71; high frequency). Some patients switched protocols during the treatment course (n = 41). All protocols involved treatment 5 days a week, with 36 treatments in a full series.

### 2.2. Depression outcome measure and data curation

The Patient Health Questionnaire - 9 Item (PHQ-9; Kroenke et al., 2001) was the primary outcome measure. PHQ-9 scores were automatically extracted from the electronic health record. Treatment and demographic information were automatically extracted, then verified manually. Discrepancies between the automated pull and manual verification were resolved via detailed review of the clinician notes by a second reviewer.

We modeled weekly PHQ-9 scores, calculated as the average of all the daily scores in a given week, with weeks defined based on the day of the week of the first treatment for each patient. Week 0 was the patient’s PHQ-9 score just before their first TMS treatment.

51 patients were excluded from analysis (see Figure 1). For patients who received fewer than six weeks of treatments (n = 9), the PHQ-9 score for their final week was carried forward. Only 13 data points out of 1666 were interpolated in this way.

**Figure 1.**
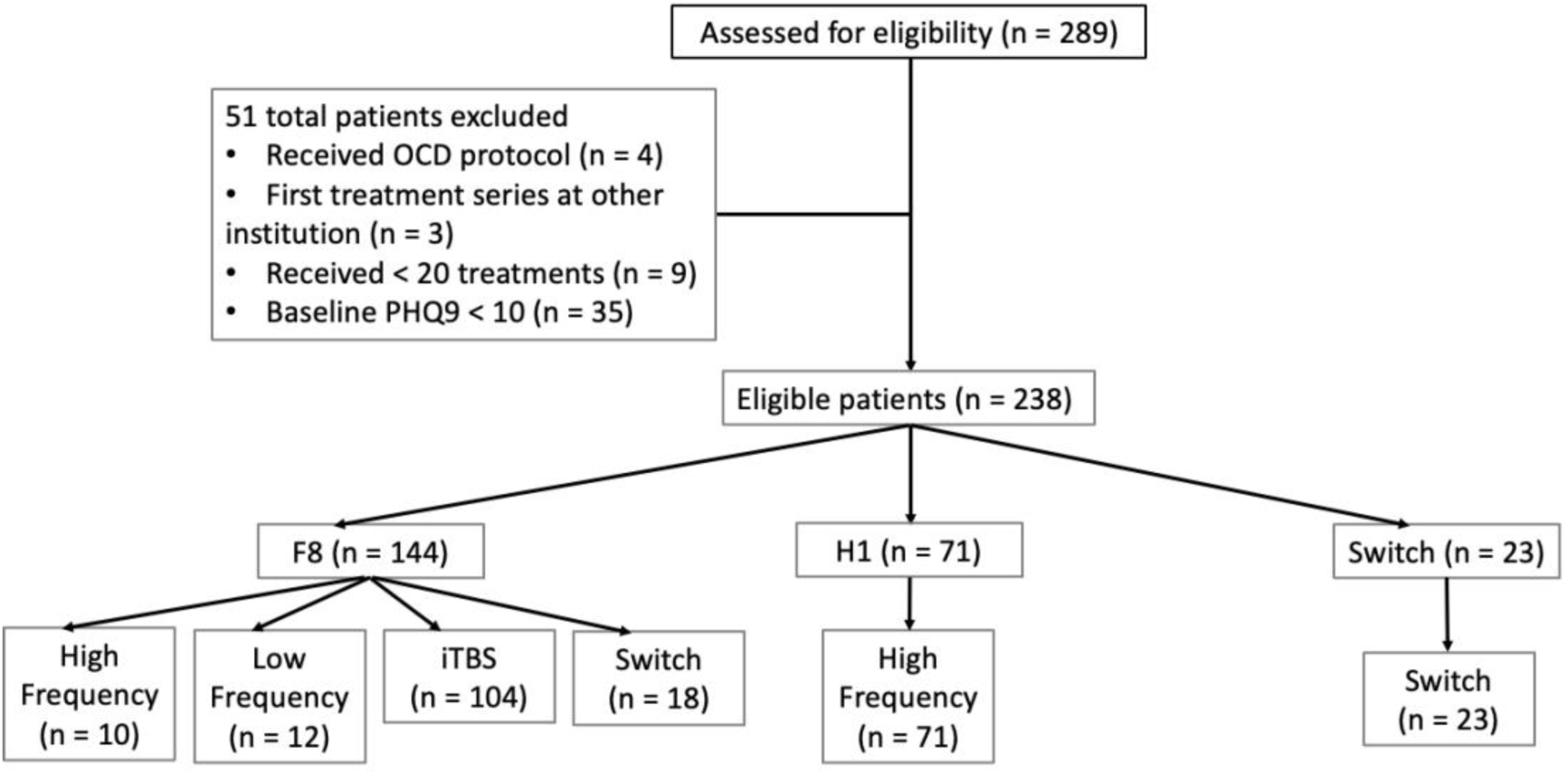
CONSORT diagram of patients included in the study. Second to last level indicates which coil patients received, last level indicates which protocol within a given coil. Switches at the protocol level indicated a protocol switch within the same coil. All coil switches were also labeled as protocol switch.

### 2.3. Trajectory modeling and model selection

#### 2.3.1. Latent class mixture modeling

Modelling was run using R (R Core Team, 2016) and scripts used to run the analyses in this report can be obtained at https://doi.org/10.5281/zenodo.11398885. LCMM used the *hlme* function in the *lcmm* package (*v2.1.0*; Proust-Lima et al., 2017). We modeled weeks 0 to 6, and tested the fit of 1 to 5 class models with linear only (L), linear and quadratic (LQ), and linear, quadratic, and cubic (LQC) polynomials. We tested model fit using 5-fold cross validation (holding out 20% of patients as a test set). Given that class assignment by LCMM can be sensitive to baseline symptom severity and that patients with different baseline symptoms may follow the same trajectory, it is possible that models may capture response trajectories better if the influence of baseline score is removed. Thus, we compared latent class models fit to raw PHQ-9 scores with models fit to baseline-corrected data. Baseline correction was performed by subtracting the baseline score from each observed PHQ-9 score.

We used multiple indicators to assess the fit of the latent class models (Chng et al., 2016; Kaster et al., 2019, 2020; Smagula et al., 2015). First, on the training data, we used BIC (Schwarz, 1978), and SABIC (Sclove, 1987), both of which perform well at selecting the correct number of classes (Tein et al., 2013).

Second, we examined how well each model generalized to new data, by examining the trained model’s predictions on the held-out test set for each fold. We predicted the scores and posterior probability of class membership for the held-out test sample. We then calculated a subject-specific trajectory by taking the weighted average of the predicted scores in each class, weighted by the posterior probability of being in that class. We assessed prediction fit with Pearson’s correlations between the observed and predicted scores.

Third, we took into account the modeled trajectories, selecting the most parsimonious model with conceptually meaningful trajectories that could be easily interpreted, and that matched known truth (Chng et al., 2016; Nylund et al., 2007). For instance, since it is clear that some TMS patients do not respond, we only accepted models that contained a nearly flat non-response trajectory. We also considered group size, rejecting models with trajectories containing less than 5% of the sample (Kaster et al., 2020, 2019; Smagula et al., 2015).

Once the optimal polynomial degree (L, LQ, LQC) and number of classes (1-5) was selected, we reran that model on the full dataset to visualize the final trajectories, and to compare descriptive and demographic information. The model using the full dataset was also used for analyses examining the factors associated with membership in the different classes (see Treatment coil and protocol effects below). Using the optimal model of the full dataset, subjects were assigned to LCMM response trajectories based on the class that yielded the highest posterior probability of class membership.

#### 2.3.2. Nonlinear mixed effects modelling

The trajectory of MDD symptom severity may also be modelled by an exponential decay function as in Equation 1 (Berlow et al., 2023):

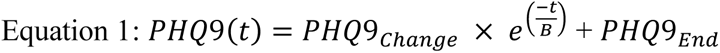

Where PHQ-9 scores at a given time point *t* are expressed as the magnitude of PHQ-9 score reduction over the course of treatment (*PHQ*9_*Change*_), which decays at a constant rate (*B*), and approaches a minimum score at the end of treatment (*PHQ*9_*End*_). NLME models (Pinheiro & Bates, 2000) were fit to the data using the *nlme* package (*v3.1-164*; Pinheiro et al., 2023), with *PHQ*9_*Change*_, *B*, and *PHQ*9_*End*_ set as group fixed effects and patient random effects. We compared the fit of this model, using BIC, and a likelihood ratio test, to a simpler model which fit only *PHQ*9_*Change*_ and *PHQ*9_*End*_ as patient random effects.

### 2.4. Predictive Power

We assessed whether trajectory models might predict response before treatment completion. We trained each of the raw LQC-4 class LCMM and baseline corrected LQC-2 class LCMM models on weeks 0-6 of the training set data, using 5**-**fold cross validation. New folds were generated independently from the previous cross validation and were stratified across trajectory classes so that each fold maintained the same proportion of subjects within each class as that of the full dataset. Using the selected model, we then predicted PHQ-9 scores and posterior probability of class membership for the test set by giving the model data for an increasing number of weeks, from week 0 to all 7 weeks of data. For the supplied weeks, we calculated subject specific predicted scores, using the weighted average described above for LCMM. For predictions of the future (hidden) weeks, we calculated a weighted average of the group level predicted scores from the training model, weighted by each test set patient’s probability of being in each class. The model-generated random effects for each patient were then added to those scores, producing patient-specific score predictions for the un-modeled weeks.

Similarly, using 5-fold cross validation, and supplying increasing weeks of data, we trained the NLME model on weeks 0-6 of training set data to estimate the time constant *B* at the population level. Then, we predicted PHQ-9 scores at the completion of treatment for the test set by rearranging Equation 1 as:

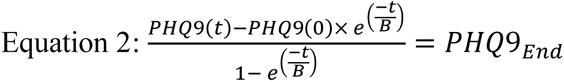

For each subject we then entered the predicted *PHQ*9_*End*_ from Equation 2 into Equation 1 to produce a predicted trajectory of PHQ-9 scores. Re-entering the predicted *PHQ*9_*End*_ into the exponential decay model allowed us to examine correlations between predicted and observed PHQ-9 scores along the entire trajectory time-series, which equates with the LCMM approach we adopted. This approach differs slightly from that employed by Berlow et al., (2023), who evaluated only the predicted value of *PHQ9_End_* as the definition of predicted response.

In each fold, we also trained the optimal LCMM model. Then, after obtaining predicted PHQ-9 scores of the test set via NLME, we entered those scores into the LCMM model and assigned participants to a predicted class based on the class that yielded the greatest posterior probability.

For both LCMM and NLME models, we tested models’ capability to predict both continuous (PHQ-9 score) and dichotomous (response) outcomes. For continuous PHQ-9 scores, we calculated the Pearson’s correlation (and 95% CI of that correlation) of observed versus predicted scores. For dichotomous (non)response, we compared modeling using two response definitions. First, we considered the traditional clinical criterion of a 50% or greater reduction in observed/predicted PHQ-9 scores at week 6. Second, after running the optimal LCMM model on the full dataset, we assigned participants to response trajectories based on the class for which the posterior probability was greatest and defined response as being assigned membership in a PHQ-9 improvement class (trajectories representing clinical improvement, but not the research definition of response). These class assignments were compared to the class assignments predicted during cross validation of the LCMM and NLME models. Consistent with best practices in biomarker research (Grzenda et al., 2021; Widge et al., 2019) we calculated sensitivity, specificity, and AUC for (non)response prediction. 95% confidence intervals were estimated in each fold using 2000 stratified bootstrap replicates. For both categorical and continuous analyses, all measures were calculated for each fold, and then averaged across folds to yield summary statistics.

### 2.5. Treatment coil and protocol effects

Multinomial logistic regression weighted by probability of class membership (Kaster et al., 2019, 2020; Smagula et al., 2015; Uher et al., 2010) was used to determine whether coil, protocol, and other patient or treatment characteristics were associated with the different trajectories.

The predictive factors tested were coil (F8 or H1), protocol (high frequency, low frequency, or iTBS), baseline PHQ-9, age, sex, any comorbid disorder, antianxiety use, and treatment year. Given that coil and protocol were highly colinear, analyses were run separately for coil and protocol. Patients who switched coils (n = 23) or protocols (n = 41) were excluded from the coil and protocol analysis, respectively. We used synthetic minority over-sampling (SMOTE; Chawla et al., 2002), as implemented in the *SmoteClassif* function in the *UBL* package (*v0.0.7*; Branco et al., 2016) to oversample the minority class and produce a balanced dataset. Data were oversampled separately for coil and protocol analyses.

Forward selection with BIC minimization (as in our prior work in Widge et al., 2016) was used to select the predictors to be included in the final model. Predictors were kept in the final model if they showed a reduction in BIC of greater than 5 on a given step (Jones et al., 2001).

## 3. Results

### 3.1. Trajectory modeling and model selection

#### 3.1.1. Latent class mixture modeling of raw PHQ-9 scores

The patient sample was overall comparable to other naturalistic, registry-based TMS studies (Carpenter et al., 2012; Sackeim et al., 2020). It was slightly female-biased and had a mean baseline PHQ-9 of 17.86. Patients may have had a higher degree of treatment resistance – mean PHQ-9 at end of treatment was 13.85, compared to 9.6 in Sackeim et al. (2020) and Carpenter et al. (2012).

There was a clear benefit for LQ over L models on BIC, SABIC, and test set prediction (Table 2). BIC, SABIC, and test set predictions also improved in the LQC models over LQ. Further, in 4 of the 5 folds, even the 3 class LQ model contained a class representing less than 5% of the population. Therefore, we selected the LQC model.

**Table 2.** Model fit statistics are used to select the final trajectory model. All values were calculated for each of the 5 folds and then averaged across folds to yield the values seen here. Changes in information criteria (ΔBIC and ΔSABIC) were calculated by subtracting the more complex model from the less complex model. Therefore, positive numbers indicate that the more complex model provided a better fit to the data. ΔBIC and ΔSABIC were calculated between the lower order polynomial and next higher order polynomial (L and LQ, LQ and LQC) within the models with the same number of classes (ΔBIC and ΔSABIC polynomial within class number). ΔBIC and ΔSABIC were also calculated between models with increasing number of classes (1 class to 2, 2 classes to 3, etc.) within the same level of polynomials (ΔBIC and ΔSABIC class number within polynomial). The models highlighted in gray were the final models selected for each approach. Models were tested independently on both raw weekly PHQ-9 scores and weekly scores that were calculated as a difference from baseline. The optimal LCMM selected was the Baseline Corrected LQC-2 model.

| Raw PHQ-9 Scores |  |  |  |  |  |  |  |  |  |
| --- | --- | --- | --- | --- | --- | --- | --- | --- | --- |
| Polynomials | Number of Classes | BIC | ΔBIC (polynomial within class number) | ΔBIC (class number within polynomial) | SABIC | ΔSABIC (polynomial within class number) | ΔSABIC (class number within polynomial) | Test set actual versus predicted score correlation | Minimum % of sample in a class |
| L | 1 | 6364.23 | NA | NA | 6345.22 | NA | NA | 0.67 | 100 |
| L | 2 | 6367.03 | NA | -2.81 | 6335.36 | NA | 9.86 | 0.67 | 42.33 |
| L | 3 | 6375.44 | NA | -8.4 | 6331.09 | NA | 4.27 | 0.66 | 11.24 |
| L | 4 | 6383.99 | NA | -8.55 | 6326.97 | NA | 4.12 | 0.66 | 8.82 |
| L | 5 | 6395.14 | NA | -11.15 | 6325.45 | NA | 1.52 | 0.63 | 5.88 |
| LQ | 1 | 6279.6 | 84.63 | NA | 6247.92 | 97.3 | NA | 0.79 | 100 |
| LQ | 2 | 6277.64 | 89.4 | 1.96 | 6230.12 | 105.24 | 17.8 | 0.79 | 16.26 |
| LQ | 3 | 6285.81 | 89.62 | -8.18 | 6222.46 | 108.63 | 7.66 | 0.77 | 6.18 |
| LQ | 4 | 6292.72 | 91.27 | -6.9 | 6213.53 | 113.44 | 8.93 | 0.75 | 6.51 |
| LQ | 5 | 6305.38 | 89.76 | -12.66 | 6210.35 | 115.1 | 3.18 | 0.74 | 3.88 |
| LQC | 1 | 6247.14 | 32.46 | NA | 6199.63 | 48.3 | NA | 0.86 | 100 |
| LQC | 2 | 6233.96 | 43.67 | 13.18 | 6167.44 | 62.68 | 32.18 | 0.85 | 11.21 |
| LQC | 3 | 6240.8 | 45.01 | -6.84 | 6155.28 | 67.18 | 12.16 | 0.84 | 10.36 |
| LQC | 4 | 6232.66 | 60.06 | 8.14 | 6128.13 | 85.4 | 27.15 | 0.83 | 11.13 |
| LQC | 5 | 6243.07 | 62.31 | -10.42 | 6119.54 | 90.81 | 8.59 | 0.81 | 5.26 |

| Baseline Corrected PHQ-9 Scores |  |  |  |  |  |  |  |  |  |
| --- | --- | --- | --- | --- | --- | --- | --- | --- | --- |
| L | 1 | 5943.45 | NA | NA | 5924.44 | NA | NA | 0.67 | 100 |
| L | 2 | 5923.04 | NA | 20.41 | 5891.36 | NA | 33.08 | 0.66 | 28.23 |
| L | 3 | 5927.15 | NA | -4.11 | 5882.8 | NA | 8.56 | 0.66 | 7.67 |
| L | 4 | 5933.12 | NA | -5.97 | 5876.1 | NA | 6.7 | 0.64 | 5.78 |
| L | 5 | 5940.75 | NA | -7.63 | 5871.07 | NA | 5.04 | 0.66 | 2.52 |
| LQ | 1 | 5690.32 | 253.13 | NA | 5658.64 | 265.8 | NA | 0.8 | 100 |
| LQ | 2 | 5663.92 | 259.12 | 26.4 | 5616.4 | 274.96 | 42.23 | 0.78 | 43.28 |
| LQ | 3 | 5672.67 | 254.48 | -8.75 | 5609.32 | 273.48 | 7.08 | 0.78 | 5.25 |
| LQ | 4 | 5687.75 | 245.37 | -15.08 | 5608.56 | 267.54 | 0.76 | 0.76 | 9.66 |
| LQ | 5 | 5703.45 | 237.3 | -15.7 | 5608.42 | 262.64 | 0.14 | 0.76 | 3.35 |
| LQC | 1 | 5560.04 | 130.27 | NA | 5512.53 | 146.11 | NA | 0.87 | 100 |
| LQC | 2 | 5493.8 | 170.12 | 66.24 | 5427.28 | 189.13 | 85.25 | 0.85 | 40.12 |
| LQC | 3 | 5504.28 | 168.4 | -10.48 | 5418.75 | 190.57 | 8.53 | 0.85 | 13.14 |
| LQC | 4 | 5521.85 | 165.9 | -17.58 | 5417.32 | 191.24 | 1.43 | 0.84 | 9.46 |
| LQC | 5 | 5535.56 | 167.89 | -13.71 | 5412.02 | 196.4 | 5.3 | 0.83 | 2.42 |

Within the LQC models, there was a slight improvement in SABIC as more classes were entered into the LQC models. For the 3 class and 5 class LQC models, BIC was slightly worse, but increased in the 2 and 4 class models. Finally, there was almost no difference in test set prediction when increasing the number of classes. Given that the BIC improvement plateaued at 4 classes, and that model is similar to that reported by another group on an independent dataset (Kaster et al., 2020), we selected the 4 class LQC model.

The final model (LQC, 4 class) was rerun on the full dataset. We labeled the resulting trajectories: lower baseline symptoms with minimal improvement (n = 112), rapid improvement (n = 38), gradual improvement (n = 25) and non-response (n = 63; Figure 2A). Patients assigned to “improvement” trajectories got better, but did not always meet the 50% threshold for response, hence the terminology. For example, only 13.39% of the patients in the minimal improvement group met the traditional 50% threshold. Two of the three improvement trajectories are not completely linear. The slope of the gradual improvement group begins shallow, and then steepens around week 2. The slope for the rapid improvement trajectory begins steeper and then levels off, indicating a good initial effect that may plateau. The lower baseline symptoms with minimal improvement group have a modest linear reduction in symptoms. We assigned this group as an “improvement” trajectory because the slope of a linear model fit to the trajectory was steeper (−0.54) than that for the non-response trajectory (−0.26), which tended to have higher baseline symptoms that remained high throughout treatment. None of those patients in the non-response trajectory met the traditional 50% reduction threshold.

**Figure 2.**
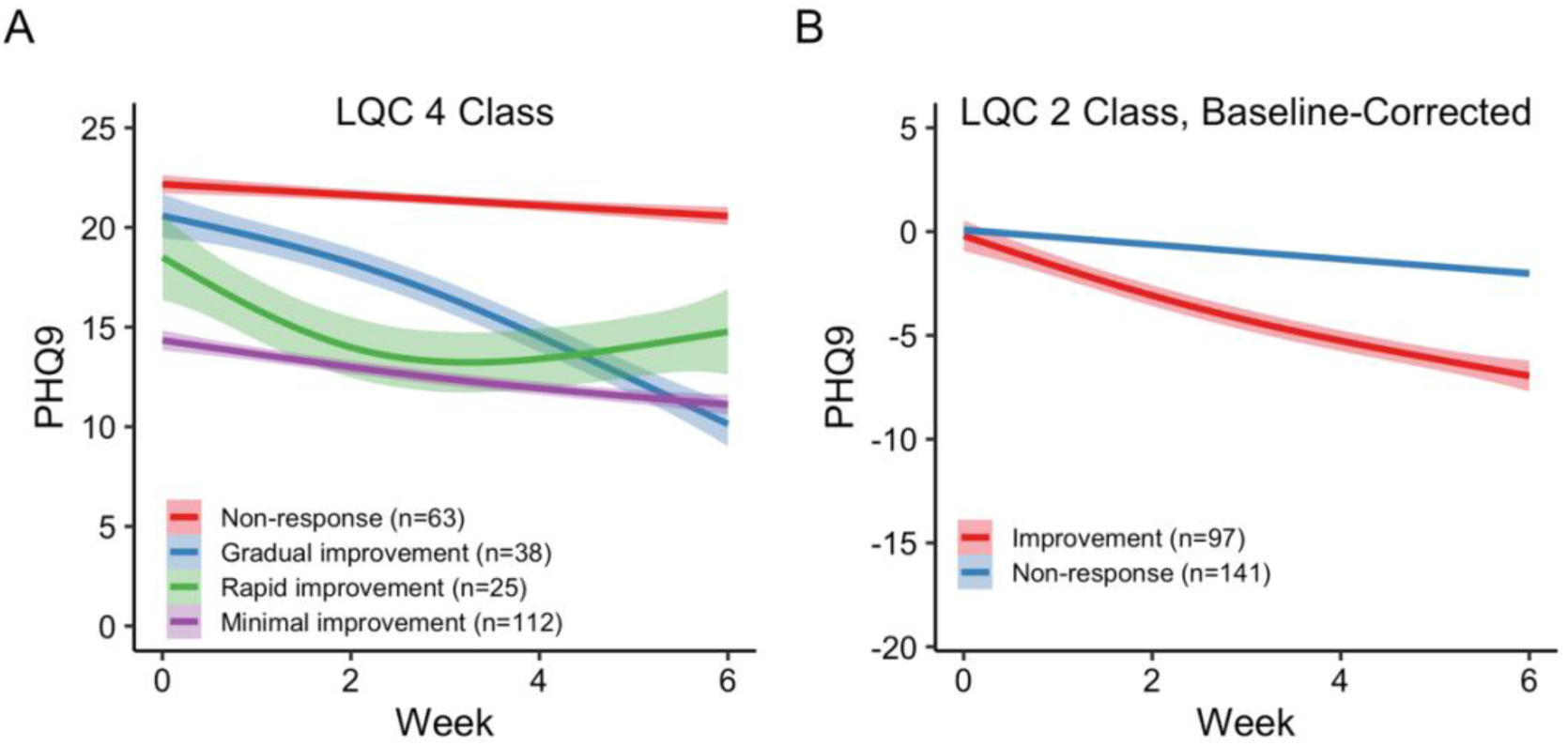
The final trajectories of the LQC 4 (A) and Baseline Corrected LQC 2 (B) class models. PHQ-9 scores are the average actual scores across the week for all the patients in each class. Error bands are SEM of the data for patients assigned to that class. *Baseline corrected LQC 2 was the final model selected and used for subsequent latent class mixture model analyses.

#### 3.1.2. Latent class modelling of difference from baseline scores

Given that LCMM can be sensitive to baseline symptom severity (i.e., PHQ-9 scores at week 0), and that patients with different baseline symptom severity may follow the same trajectory, it is possible that models may capture response trajectories better if the influence of baseline score is removed. Thus, we ran additional LCMMs with PHQ-9 scores computed as a difference from baseline. Similar to raw PHQ-9 scores, BIC, SABIC, and test set predictions improved as quadratic and cubic polynomials were added to the model (Table 2). Thus, we selected the LQC model for baseline corrected scores.

Within the LQC models, BIC improved when the number of classes was increased from 1 to 2. BIC did not improve any further with the addition of more classes. SABIC improved as more classes were included, but the improvement of SABIC was modest after 2 LQC classes.

Test set predictions showed little change as classes were added to the models. Given the plateau of BIC and SABIC improvement at 2 classes, and little change in test set prediction, we settled on the 2 class LQC model for baseline corrected PHQ-9 scores.

We ran the final model of baseline corrected PHQ-9 scores (LQC, 2 class) on the full dataset, labelling the resulting trajectories: **improvement** (n = 97), and **non-response** (n = 141; Figure 2B). The patients assigned to the “improvement” trajectory had a mean (SD) reduction in PHQ-9 scores of −6.87 (6.03), but did not always meet the traditional 50% criterion for response – only 41.23% of patients in the improvement trajectory met this criterion. Patients assigned to the non-response trajectory showed a smaller reduction in PHQ-9 scores at week 6 of treatment (M (SD) = −2.05 (2.52)), and only 2.84% of those patients met the traditional 50% criterion for response. Table 1 contains the patient characteristics for each trajectory. Overall, correlations between test set predictions and observed data were largely comparable between the raw PHQ-9 LCMM model and the baseline corrected one. However, within polynomials in the baseline-corrected model, there were clear improvement in both BIC (range = 5504.28 - 5943.45) and SABIC (5412.02 - 5924.44) in comparison to the model fit to raw PHQ-9 scores (BIC range = 6232.66 - 6395.14; SABIC range = 6119.54 - 6345.22). Therefore, we selected the baseline-corrected LQC 2 class model as the optimal LCMM.

#### 3.1.3. Nonlinear mixed effects modelling of PHQ-9 scores

We fit the exponential decay function to data using NLME models which included *PHQ*9_*Change*_, *B*, and *PHQ*9_*End*_ as random effects at the patient level. This model yielded an AIC = 8042.47, BIC = 8097.05, and a log likelihood of −4011.43. These values indicated a better fit than a simpler model which included only *PHQ*9_*Change*_ and *PHQ*9_*End*_ as random effects, with an AIC = 8357.46, BIC = 8395.38, and log likelihood of −4171.73. The more complex model also yielded a significant likelihood ratio test over the simpler one, LRT = 320.59, *p* < .001. Thus, we included all random effects in the final nonlinear mixed model.

### 3.2. Predictive Power

Trajectory modeling using LCMM had acceptable predictive power (Table 3). When given raw PHQ-9 scores for week 0 (and up), the correlation between predicted and observed scores was high. When response was defined by assignment to an improvement trajectory, the AUC exceeded chance at week 0 for raw PHQ-9 scores. These results are likely due to the large contribution of baseline PHQ-9 to final trajectory in the raw score modeling approach. When response was defined as a >50% reduction in PHQ-9, the AUC did not exceed chance until the model was given raw PHQ-9 scores for weeks 0-4. Specificity was high, starting at week 0, but sensitivity improved when given data for an increasing number of weeks. This indicates that the model is biased to predict non-response initially, becoming less so when given more weeks of data.

**Table 3.**
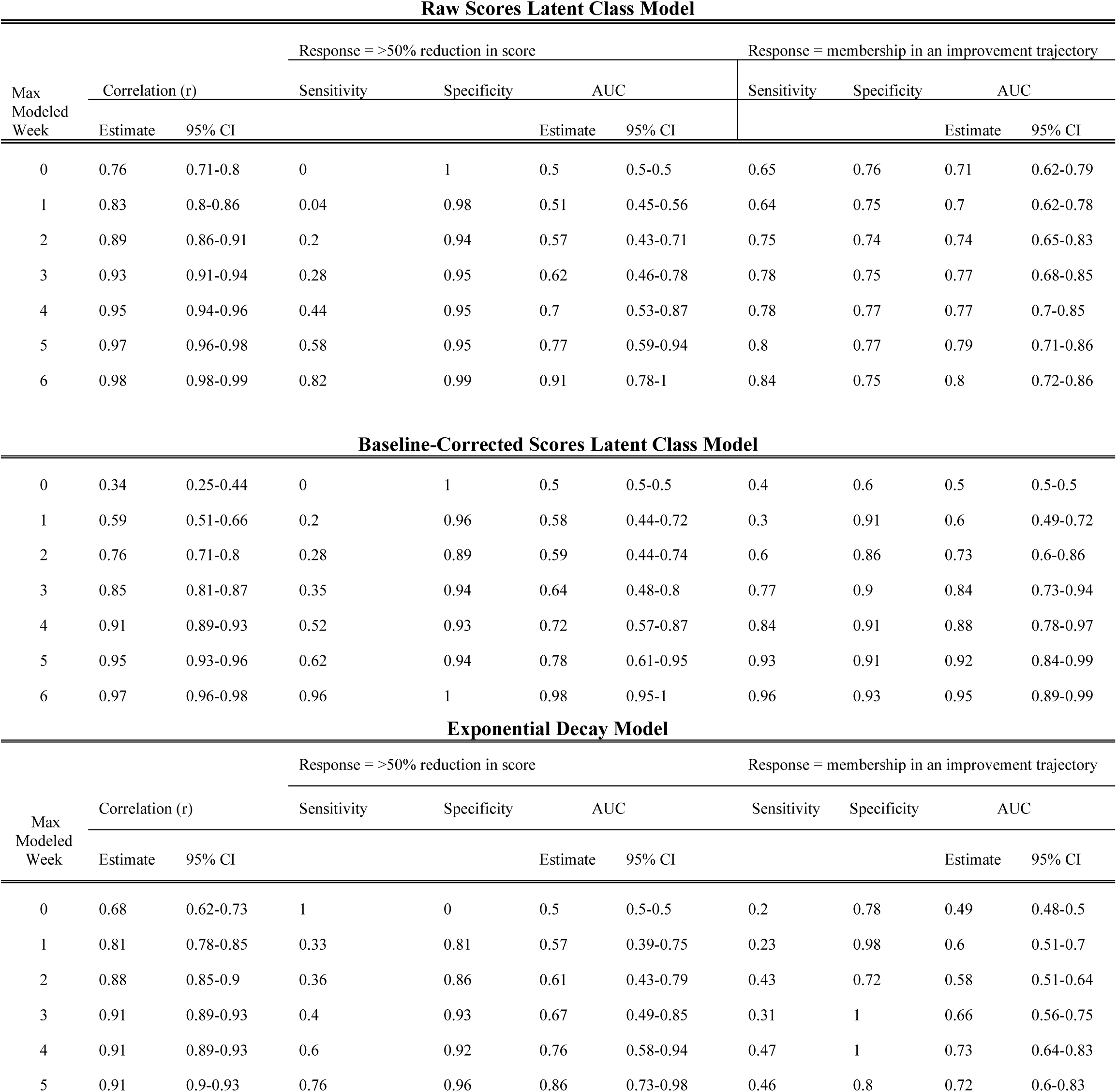
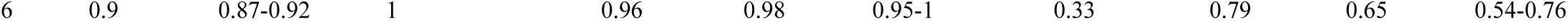
Predicting response with trajectory modeling. Max modeled week indicates the maximum week number of test set data that was given to the model to make predictions, up to 6 (treatment completion). Pearson correlations (r) and 95% confidence intervals compare the predicted versus observed scores of the test set. Sensitivity, specificity and AUC quantify categorical (non)response prediction when response was defined as: 1) the traditional method of 50% or greater reduction in score, and 2) assigned membership in the improvement trajectory of the latent class mixture model of baseline corrected scores.

When removing the influence of baseline scores on LCMM trajectories, correlations of predicted with observed scores were generally lower than those of raw scores, particularly when 0 or 1 weeks of data were provided. In comparison to raw scores, the AUC of predicting a 50% PHQ-9 score reduction was greater when given just one week of data, and remained consistently higher as more weeks of baseline-corrected data were entered. However, AUC did not exceed chance until 0-4 weeks of data were entered, for both the raw and baseline-corrected score models. When predicting improvement trajectory assignment, AUC was lower for baseline corrected scores when only the first two weeks of data were entered. This is again likely due to the influence of baseline scores on trajectory assignment when modelling raw PHQ-9 scores. However, AUC improved beyond raw scores when three or more weeks of baseline corrected data were entered.

The NLME approach also yielded acceptable predictive power. Across the five folds of training, estimates of *B* at the population level had a mean (SD) = 2.11 (0.09). Predicted values of *PHQ*9_*End*_ on test sets were highly correlated with observed values, *r* [95% CI] = .71[.68 - .73], *p* < .001. Correlations of observed PHQ-9 scores to predicted scores using the exponential decay function were generally lower than those obtained from the LCMM approach. AUC for predicting a 50% score reduction was numerically higher for NLME than for LCMM-based models. However, similar to the LCMM models, AUC did not exceed chance until weeks 0-4 were entered into the model. When response was defined as assignment to an LCMM improvement trajectory based on predicted scores, AUC was lower in comparison to the LCMM models, even though the NLME model reached significance (AUC confidence interval excluding 0.5) at week 1 compared to week 2 for the baseline-corrected LCMM. This suggests that the exponential decay model is useful at predicting larger reductions in PHQ-9 scores during treatment, but less so when symptom improvement is smaller and does not meet the traditional 50% reduction criterion.

**Figure 3.**
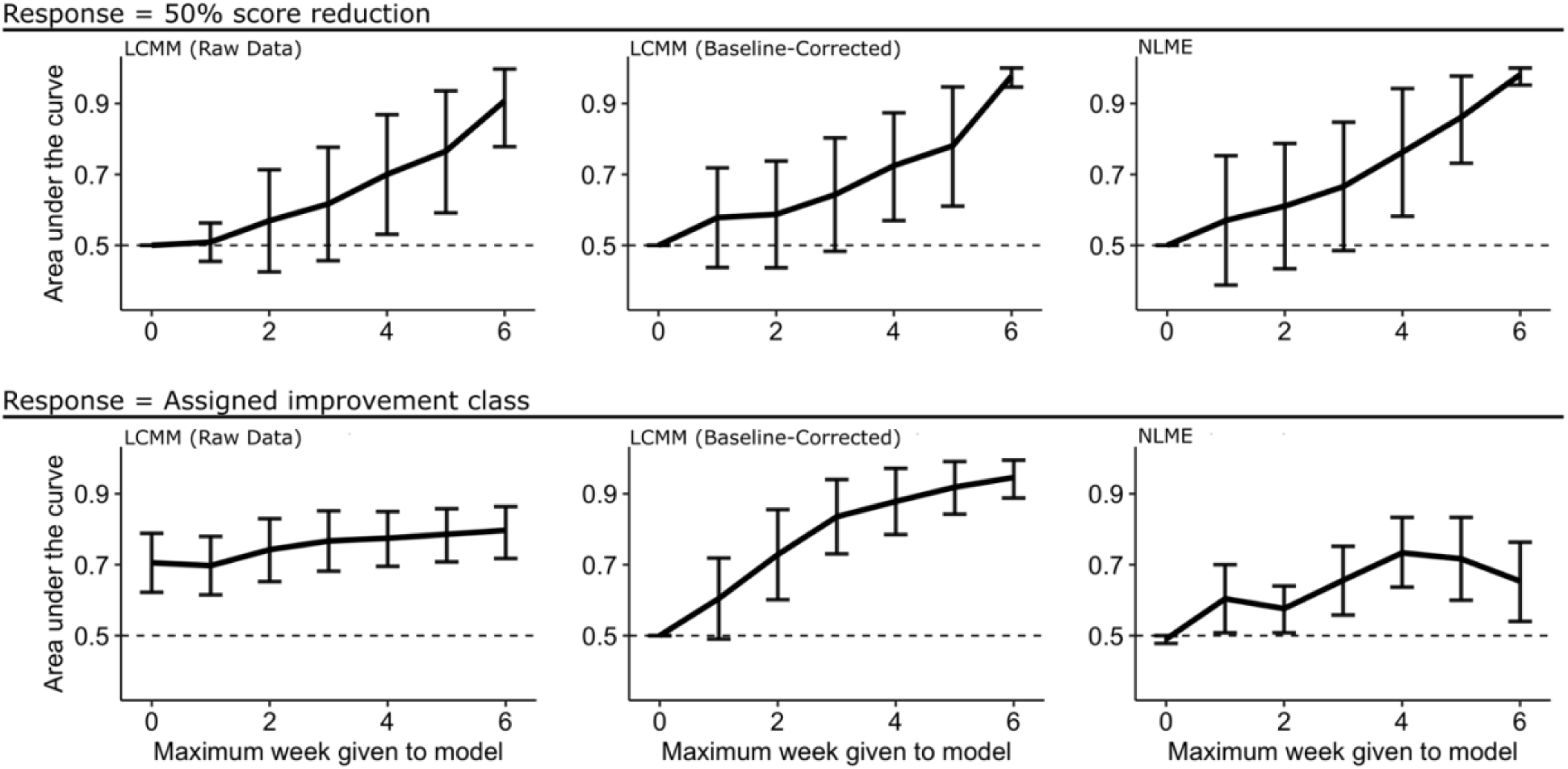
Area under the receiver operating characteristic curve for model predictions of treatment outcomes with increasing observations. The dashed line in each plot shows chance (AUC = 0.5), and error bars denote the 95% CI of each estimate (see Table 3). Top row shows AUC for prediction of traditional definition of response (50% PHQ-9 score reduction). Bottom row shows AUC for prediction of assignment to the LCMM improvement trajectory. Left column shows prediction results for LCMM of raw data. Middle row shows prediction results for LCMM of baseline-corrected data. Right column shows prediction results for NLME.

### 3.3. Treatment coil and protocol effects

None of baseline PHQ-9, age, sex, treatment year, or antianxiety use differed between patients treated with the F8 or H1 coil. Both coil and protocol increased BIC from the null model in logistic regressions of trajectory class assignment. The stepwise addition of baseline PHQ-9, age, sex, treatment year, and antianxiety medication use increased BIC from the null model in each case. Thus, in this dataset, neither standard clinical factors nor treatment coil influenced the overall probability of response.

## 4. Discussion

We modeled trajectories of TMS response in a naturalistic sample, demonstrating that prior results in well characterized clinical trial samples can generalize to more heterogeneous populations. When modelling raw PHQ-9 scores, a linear-quadratic-cubic, four class latent mixture model provided the best fit while also being the most parsimonious. Those trajectories largely replicated prior trajectory modeling in TMS (Kaster et al., 2020, 2019). However, we found that baseline scores may have a large influence on the classes that are derived, while providing little information on the final symptom trajectory. For example, patients with different baseline symptom severity, but following the same trajectory, may be incorrectly dichotomized into separate response classes. In support of this, a latent class model that accounted for baseline symptom severity yielded a superior fit to data, along with greater performance in predicting treatment response. The optimal latent class model for those data was a linear-quadratic-cubic 2 class model. This suggests that when accounting for baseline symptom differences, there are two nonlinear trajectories underlying response to TMS – those who receive benefit and those who do not. This contrasts previous suggestions that antidepressant response trajectories reveal subgroups of responders that may be biologically separable (Kaster et al., 2020, 2019).

In addition, we compared a separate approach which defines antidepressant symptom trajectories as an exponential decay function (Berlow et al., 2023). This approach does not impose explicit subgroups on response trajectories, but rather, denotes response as an individual’s magnitude, rate, and outcome of treatment response. Here also, our results partially replicated and partially diverged from the original report. We confirm the exponential model’s predictive power, and show that it can exceed categorical trajectory models in some situations (numerically higher AUCs when predicting 50% drop in PHQ-9). We did not replicate the original finding that exponential models can predict response as soon as week 1; in our dataset, prediction above chance was only possible from week 4 onward. Similarly, the exponential decay approach substantially underperformed the latent class (categorical) model when predicting assignment to an overall improvement trajectory, at all modeled timepoints. This may reflect the fact that the trajectory concept is built in to the latent class approach, and thus these models are more able to consider small symptom improvements that may be meaningful to the patient, but do not reach traditional response definitions.

A larger point is that both models can predict clinical outcomes before completion of the TMS course. In predicting the traditional criterion of a 50% score reduction, all models we evaluated could predict response better than chance after 4 weeks of treatment. When predicting assignment to an “improvement” trajectory, the latent class model of raw PHQ-9 scores exceeded chance when given only baseline scores. However, with increasing weeks of data, predictive performance of this model showed little improvement. This suggests that baseline scores are overly-influential in latent class models and that those scores provide little benefit in defining the final symptom trajectory. Rather, the predictive performance of the baseline corrected latent class model suggests that baseline scores should be accounted for when modelling antidepressant response trajectories to TMS with latent class mixture models. When accounting for baseline symptom severity, the latent class model could predict assignment to a (non)improvement trajectory when given data from only 2 weeks of treatment.

The clearest clinical use of trajectory modeling would be identifying (non)responders without needing to provide a full 6-week treatment series. Using a standard definition (>50% improvement), all models we evaluated could predict response better than chance after 4 weeks of treatment. This may be of limited practical value; with two thirds of the treatment course completed, there is only modest cost savings from identifying non-responders. Response prediction at 4 weeks may be more useful if conclusive evidence emerges that a coil or protocol switch can “rescue” a patient into response. This is a strong anecdotal belief among TMS clinicians, but has not been empirically validated. Using simpler modeling, researchers have shown that symptom improvement at 10 sessions is predictive of ultimate clinical response to TMS (Feffer et al., 2018), suggesting the possibility of a more optimal method of predicting response early in treatment. Importantly, although both latent class and exponential models had only modest predictive power, they far exceeded the predictive power of easily observed factors such as concomitant medication, age/sex, or type of treatment coil used. This highlights that, in general, such modeling is valuable. Another potential value of response trajectory models might lie in biomarker discovery. Although the models have modest predictive power to predict the future from clinical data alone, the trajectories/exponential parameters remove some of the noise present in raw mood ratings from the PHQ-9. By providing a lower-noise dependent variable, trajectory models might enable discovery of baseline or treatment-emergent physiologic markers to alter TMS treatment. Similar denoising approaches have led to the discovery of new biomarkers for invasive brain stimulation (Basu et al., 2023; Sani et al., 2018).

There are several limitations in our study, mainly due to the naturalistic sample. For example, many patients were treated with multiple coils, or with multiple protocols. While we included these patients, it is critical to understand how these switches may alter outcomes. Our current dataset contains too few observations of each switch type to reliably perform this analysis. Further, switches occurred based on a subjective clinician recommendation, and at variable time points. The present research provides evidence for a more systematic means by which clinicians can determine whether a coil or protocol switch might be warranted. Further, we believe that the “messiness” inherent in a naturalistic sample is also a strength, given that the long-term goal of this type of modeling is for it to be used in clinical practice.

In summary, we have demonstrated that previously suggested subgroups of antidepressant response to TMS may be better defined as simple response/non-response trajectories. We compared two approaches of modelling response trajectories and show that treatment response can be reliably predicted given 4 weeks of treatment data, and symptom severity changes along an improvement trajectory can be predicted given two weeks of treatment data. Thus, we provide preliminary evidence that trajectory modeling might be viable for guiding treatment decisions. This would require a clearer understanding of reasonable stepped-therapy TMS algorithms, an area of active research. It is not yet clear if similar modeling could guide the use of more compressed TMS protocols, e.g. if these same trajectories might appear during a course of TMS given over a week (Cole et al., 2020). Longitudinally measured biomarkers should show similar trajectories that precede the clinical response, which would provide strong evidence of causality.

## Acknowledgments

We thank the University of Minnesota Biomedical Informatics and Data Access (BPIC) for their work with data extraction and curation.

## Author Contributions

ANM: Formal analysis, Data Curation, Writing - Revisions, Visualization, Conceptualization; STO: Formal analysis, Data Curation, Writing - Original Draft, Visualization, Conceptualization; CRPS: Project administration, Data Curation; DCC: Data Curation; SW: Data Curation, Resources; AIS: Data Curation; SCA: Resources, Supervision; SCO: Resources; CBP: Resources; BRR: Resources; AH: Resources; MB: Resources; ZN: Resources, Supervision; ASW: Conceptualization, Funding acquisition, Supervision, Resources; All authors participated in Writing - Review & Editing

## Data availability statement

Deidentified data will be made available upon request.

